# Association Between Preoperative Myocardial Perfusion Imaging and Cardiac Events after Elective Noncardiac Surgery

**DOI:** 10.1101/2023.07.10.23292481

**Authors:** Seong-Bong Wee, Cheol Hyun Lee, Tae Joon Jun, Jung-Min Ahn, Jeong Hwan Yook, Jae-Sik Nam, In-Cheol Choi, Dae Hyuk Moon, Sangwon Han, Hoyun Kim, Yeonwoo Choi, Jinho Lee, Sangyong Cho, Tae Oh Kim, Do-Yoon Kang, Pil Hyung Lee, Duk-Woo Park, Soo-Jin Kang, Seung-Whan Lee, Young-Hak Kim, Cheol Whan Lee, Seong-Wook Park, David J Cohen, Seung-Jung Park

## Abstract

**Background:** There remains a lack of robust evidence regarding the prognostic value of myocardial perfusion imaging (MPI) before noncardiac surgery in large and diverse patient populations.

**Methods:** This retrospective observational cohort study from single, tertiary, high surgical volume center in South Korea included 82,441 patients aged >40 years who underwent MPI using pharmacologic stress single photon emission computed tomography within 6 months before elective noncardiac surgery from January 2000 to December 2021. Results of MPI were classified as abnormal (any fixed or reversible perfusion defect) vs normal MPI before noncardiac surgery. The primary outcome was a composite of cardiac death or myocardial infarction within 30 days.

**Results:** Among the 82441 patients (mean±standard deviation age, 65.7±9.6 years; 47417 [57.5%] men), 184 (0.2%) experienced cardiac death or myocardial infarction within 30 days after noncardiac surgery. MPI were abnormal in 5603 patients (6.8%). Compared with a normal MPI, an abnormal MPI had a higher risk of the primary outcome (crude incidence, 1.2% vs 0.1%; adjusted odds ratio, 4.64; 95% confidence interval, 3.29-6.50; *P*<.001). The presence of an abnormal MPI improved discrimination for the primary outcome (area under the receiver operating characteristic curve with MPI vs without MPI (0.77 vs 0.73; *P*<0.001)) and significantly increased net reclassification improvement (0.26; 95% confidence interval, 0.11-0.40; *P*<.001). Among patients with an abnormal MPI, 378 (6.7%) underwent pre-operative coronary revascularization; however, this was not associated with a lower risk of the primary outcome (*P*=.56).

**Conclusion:** An abnormal myocardial perfusion imaging appeared to be an important risk factor for adverse postoperative events and provided additional prognostic value for patients undergoing noncardiac surgery. Nevertheless, preoperative MPI was limited by its low positive predictive value for postoperative cardiac events, leading to potentially unnecessary coronary revascularization procedures with unproven prognostic value.

**Clinical Perspectives:** *What is new?:*

This study included the largest population to date and included a broad spectrum of patient and surgical procedures compared with previous studies limited by relatively small sample sizes and low event rates.
Preoperative abnormal MPI results were significantly associated with the postoperative risk of cardiac death and myocardial infarction in patients undergoing noncardiac surgery, and provided incremental prognostic value beyond clincial assement only.

*What are the clinical implications?:*

- ur study suggested that MPI appears warranted, particularly for patients with a considerable surgical risk.
- However, its low positive predictive value, and unproven prognostic value of coronary revascularization triggered by MPI should be taken into account in the clincial practice.

## Introduction

Every year, ≥200 million adults undergo major noncardiac surgery and its number is still increasing.^1, 2^ Despite the overall safety of contemporary noncardiac surgery, approximately 10% of these patients experience post-operative complications.^3^ Cardiovascular complications remain the leading cause of death within 30 days of noncardiac surgery.^4^ Therefore, identification of patients at high cardiovascular risk during preoperative consultation is important.

Previous studies have revealed that abnormal features upon myocardial perfusion imaging (MPI) indicate an increased risk of perioperative cardiac complications.^5^ Current practice guidelines recommend stress MPI prior to non-cardiac surgery for patients with both elevated risk of major adverse cardiac events and poor functional capacity especially if testing impacts decision-making or perioperative care.^4^ The uncertain value of pre-operative MPI derives from its low diagnostic yield, the unclear clinical benefit of preoperative revascularization triggered by its results, and the potential for unnecessary delays of surgical treatment.^6, 7^ Notably, previous studies on preoperative MPI were limited by their small samples and numbers of events.^8^ Most studies were performed on the highest-risk patients (eg, those undergoing vascular surgery) decades ago, and the application of those results in today’s practice is unclear, given advances in both surgery and perioperative care. In addition, the predictive discrimination associated with MPI has not been adequately compared with those derived from preoperative risk calculators alone.^9^ Nevertheless, preoperative MPI and subsequent revascularization are frequently performed in real-world practice to evaluate cardiac risk in an effort to prevent perioperative cardiac complications.^10–12^

To address these gaps in contemporary evidence, we performed a retrospective, real-world study: (1) determine the prognostic value of preoperative MPI to predict cardiac events after elective noncardiac surgery; and (2) examine the clinical benefit of selective coronary angiography and revascularization in response to abnormal MPI.

## Methods

### Study Design and Study Population

This was a single-center, retrospective observational cohort study, and was conducted using data from the Asan Biomedical Research Environment (ABLE), which is a de-identified clinical database of Asan Medical Center, a 2700-bed tertiary hospital in Seoul, South Korea. This data warehouse contains all medical records of our center, including electronic medical records, international classification of disease codes, laboratory findings, imaging data, and medications in an anonymized form.^13^

The study population was drawn from all patients who underwent MPI in the 6 months prior to elective noncardiac surgery under general anesthesia between January 2000 and December 2021. Patients were excluded if they met any of the following criteria: younger than 40 years of age; undergoing an emergency operation; experiencing acute myocardial infarction in the month before surgery; undergoing cardiac surgery; undergoing nonsurgical procedures (eg, bronchoscopy, endoscopy, cystoscopy, and percutaneous vascular or nonvascular procedures); undergoing minor surgery with minimal sedation or local anesthesia, such as skin, dental, and ophthalmologic procedures. Only the index procedure of patients undergoing multiple eligible procedures during the study period was used for analyses. Additional inclusion and exclusion criteria are summarized in the Supplement Appendix.

This study conformed to the ethical guidelines of the Declaration of Helsinki and was approved by the institutional review board of Asan Medical Center. The need for written informed consent was waived. No industry was involved in the design, conduct, or analysis of the study.

### Data Extraction and Collection

Patient demographics, comorbidities, prescriptions, laboratory data, types of surgeries, and outcomes were obtained via the ABLE system by researchers who were blinded to the process of data analysis. Comorbidities diagnosed prior to the date of noncardiac surgery were electronically obtained using the Korean Standard Classification of Diseases and Causes of Death (KCD-7), which was developed based on the International Classification of Diseases, 10th revision (Supplement).^14^ In addition, the revised cardiac risk index (RCRI), which consists of six identifiable predictive factors (high-risk surgery [intraperitoneal, intrathoracic, and suprainguinal vascular surgery], ischemic heart disease, congestive heart failure, cerebrovascular disease, diabetes mellitus controlled with insulin therapy, and renal dysfunction [serum creatinine concentration >2.0 mg/dL]), was calculated. All 1436 types of surgeries performed in the study population were reviewed and classified as low- or high-risk surgeries based on prior expert consensus (Supplement Appendix).^15, 16^

### Myocardial Perfusion Imaging

Single photon emission computed tomography with thallium-201 (Tl-201) was used to acquire myocardial perfusion images via a standardized protocol, as previously described.^17^ Pharmacologic stress was induced with intravenous infusion of either adenosine (0.14 mg/kg/min for 6 min) or dipyridamole (0.56 mg/kg/min for 4 min). At peak stress, a 44.4– 148.0-MBq dose of 201-Tl was intravenously injected, depending on the patient’s body weight and the type of gamma camera used. Post-stress and redistribution MPIs were acquired with one of the following camera systems equipped with a conventional Anger camera or cadmium-zinc-telluride detectors: Triad 88 or XLT (Trionix Research Laboratory, Twinsberg, OH, USA); ADAC or Precedence 16 (Philips Healthcare, Best, The Netherlands); E.Cam, Symbia T2, or Evo Excel (Siemens Healthineers, Erlangen, Germany); Infinia, Ventri, Discovery NM830, or NM530c (GE Healthcare, Waukesha, WI, USA). Specific acquisition parameters depended on the type of camera.

MPI was primarily analyzed qualitatively by experienced nuclear medicine physicians (D.H.M. and S.W.H) as a normal or abnormal.^17^ Subsequently, abnormal results were further classified into fixed perfusion defects only or any reversible perfusion defect. In addition, semi-quantitative analysis, performed using a 20-segment model and a five-point scale, was used to calculate the summed stress score, summed rest score, and summed difference score (SDS). The SDS was converted into a percentage of total myocardium by dividing it by the maximum potential score (4 × 20) to assess the ischemic burden (% ischemic myocardium).^18, 19^

### Study Outcomes and Follow-up

The primary outcome in this study was the composite of cardiac death or myocardial infarction within 30 days after elective noncardiac surgery. Cardiac death was defined as sudden death or death secondary to a proximate cardiac cause, including cardiac arrest, myocardial infarction, low-output failure, or fatal arrhythmia. Myocardial infarction was defined as an elevation of cardiac enzymes with associated signs and symptoms of ischemia felt to be caused by coronary atherothrombosis. The secondary outcomes were cardiac death, all-cause death, and myocardial infarction within 30 days after elective noncardiac surgery. The mortality data was confirmed by cross-referencing with the Korean National Health Insurance Service, which is a single-payer program of a universal health coverage system and mandatory health care in Korea.^20^ In addition, all medical records and other source documents were carefully reviewed, by two physicians (S.B.W. and C.H.L), blinded to MPI results, to validate the diagnosis of cardiac death and myocardial infarction. Myocardial injury after noncardiac surgery (MINS) was defined as a postoperative cardiac troponin concentration above the 99^th^ percentile of the upper reference limit of the assay without evidence of nonischemic etiology among patients who underwent a routine troponin test after noncardiac surgery.^21^ Definitions of the study outcomes are provided in the Supplement Appendix.

## Statistical Analysis

Baseline characteristics of the patients are reported as frequencies and percentages for categorical variables and means with standard deviations for continuous variables. Survival was assessed using the Kaplan–Meier method and compared using the log-rank test. We compared the primary and secondary outcomes according to the MPI results by using logistic regression models, and the final multivariable models included age, sex, the RCRI, and MPI results. These covariates were selected a priori based on previous evidence.^22^ Odds ratios (ORs) and corresponding 95% confidence intervals (CIs) were reported. We assessed the risk prediction and stratification performance of MPI by calculating the area under the time-dependent receiver operating characteristic (ROC) curve and the continuous net reclassification improvement (NRI). Because of the potential for type I error due to multiple comparisons, for which we did not adjust for the *P* values, results of analyses for secondary outcomes should be interpreted as exploratory. All reported *P* values are two-sided. A *P* value <.05 was considered statistically significant. Analyses were performed using R software, version 4.2.1 (R Foundation for Statistical Computing, Vienna, Austria).

## Results

### Characteristics of the Population

From January 2000 to December 2021, 82,441 patients who underwent MPI for preoperative cardiac risk assessment before elective noncardiac surgery were included in this study, of whom 5603 (6.8%) had an abnormal MPI (Figure 1). The patients’ mean age was 65.7±9.6 years, 57.5% were men, 50.2% underwent high-risk surgery, and 12.2% had an RCRI score ≥2 (Table 1). Compared with patients with normal MPI, patients with abnormal MPI were more likely to have comorbidities. The frequency of abnormal MPI increased as the RCRI score increased, ranging from 2.4% among patients with an RCRI 0 to 50.3% in patients with RCRI ≥4 (Figure 2).

**Figure 1.**
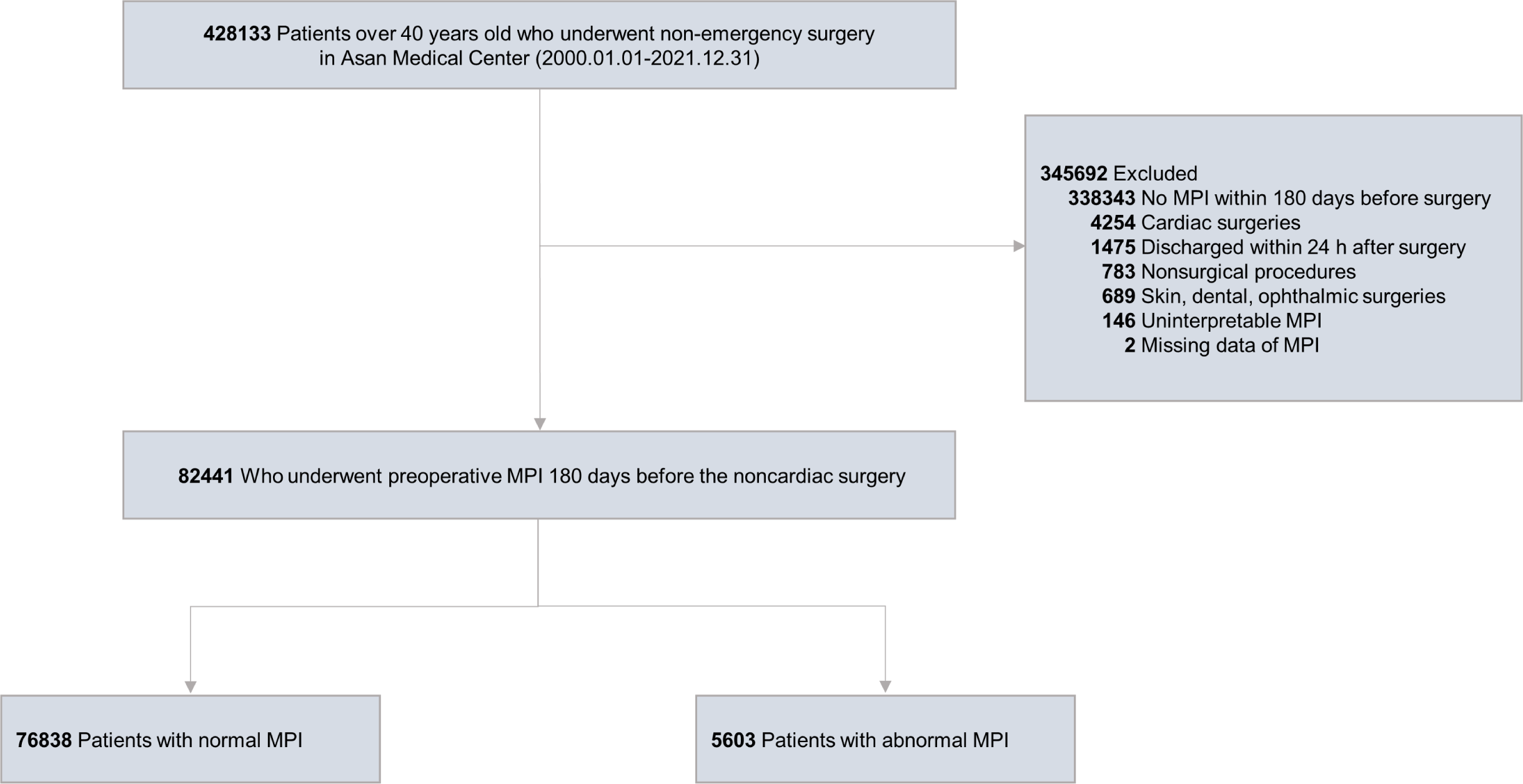
Flow Diagram of Participants in the Study. MPI, myocardial perfusion imaging.

**Figure 2.**
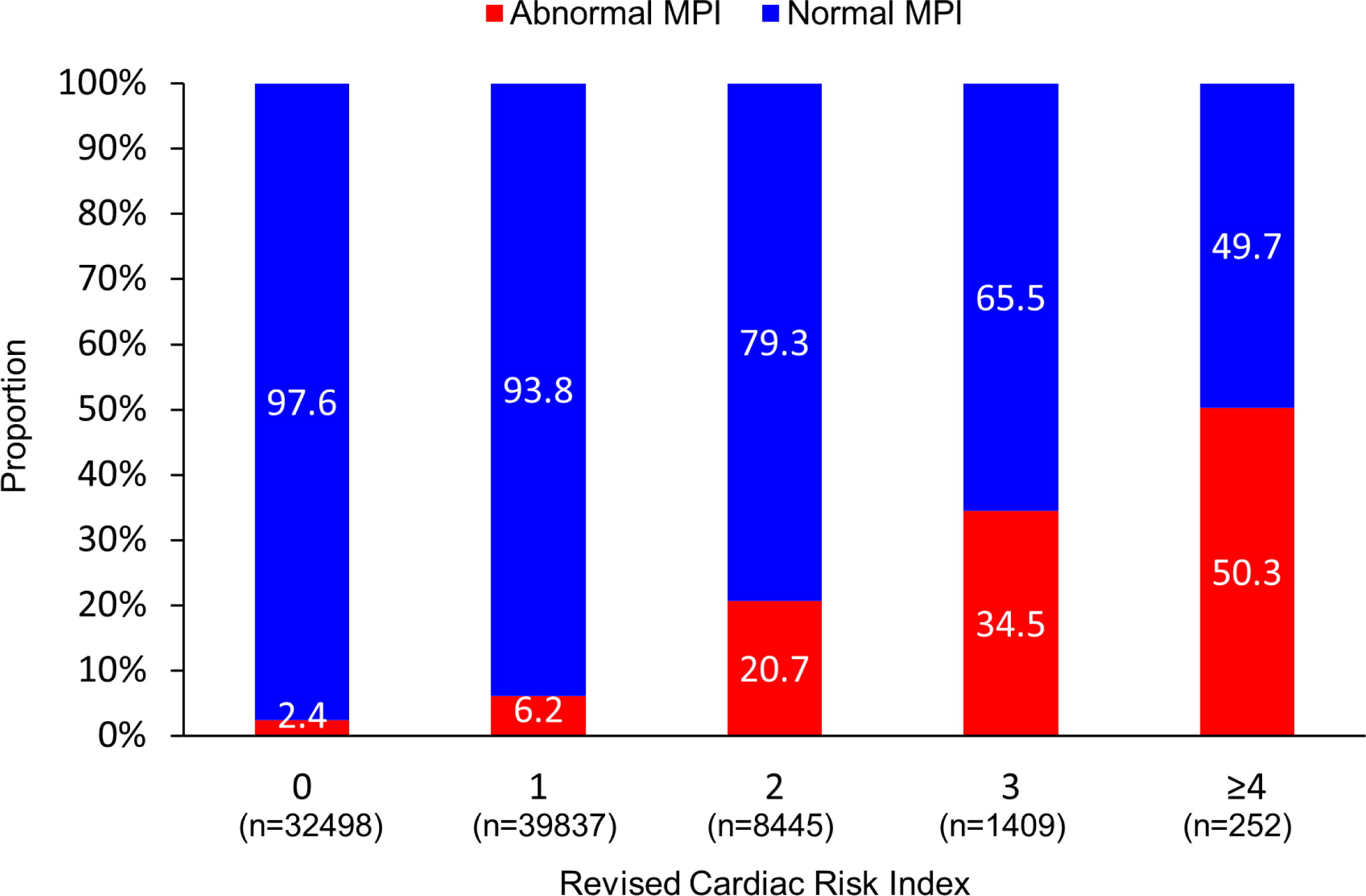
Frequency of Abnormal MPI According to the Revised Cardiac Risk Index. MPI, myocardial perfusion imaging.

**Table 1.**
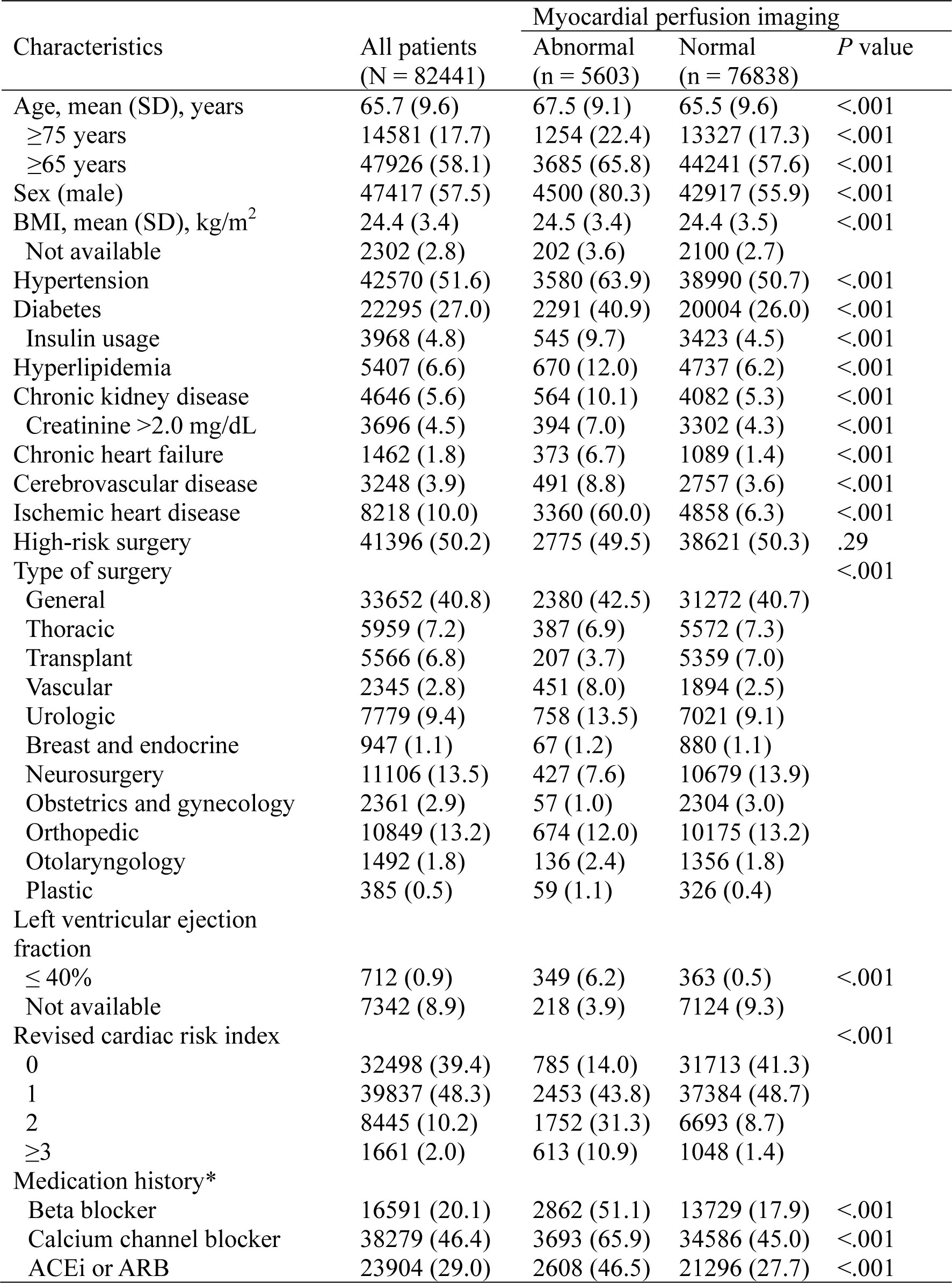
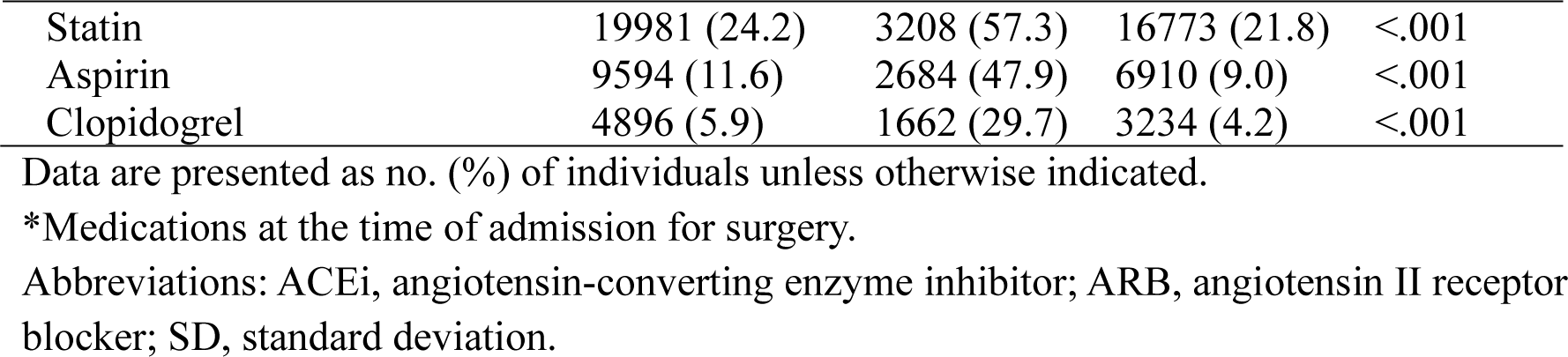
Baseline Characteristics.

| Characteristics | All patients<br>(N = 82441) | Myocardial perfusion imaging |  | P value |
| --- | --- | --- | --- | --- |
|  |  | Abnormal<br>(n = 5603) | Normal<br>(n = 76838) |  |
| Age, mean (SD), years | 65.7 (9.6) | 67.5 (9.1) | 65.5 (9.6) | <.001 |
| ≥75 years | 14581 (17.7) | 1254 (22.4) | 13327 (17.3) | <.001 |
| ≥65 years | 47926 (58.1) | 3685 (65.8) | 44241 (57.6) | <.001 |
| Sex (male) | 47417 (57.5) | 4500 (80.3) | 42917 (55.9) | <.001 |
| BMI, mean (SD), kg/m<sup>2</sup> | 24.4 (3.4) | 24.5 (3.4) | 24.4 (3.5) | <.001 |
| Not available | 2302 (2.8) | 202 (3.6) | 2100 (2.7) |  |
| Hypertension | 42570 (51.6) | 3580 (63.9) | 38990 (50.7) | <.001 |
| Diabetes | 22295 (27.0) | 2291 (40.9) | 20004 (26.0) | <.001 |
| Insulin usage | 3968 (4.8) | 545 (9.7) | 3423 (4.5) | <.001 |
| Hyperlipidemia | 5407 (6.6) | 670 (12.0) | 4737 (6.2) | <.001 |
| Chronic kidney disease | 4646 (5.6) | 564 (10.1) | 4082 (5.3) | <.001 |
| Creatinine >2.0 mg/dL | 3696 (4.5) | 394 (7.0) | 3302 (4.3) | <.001 |
| Chronic heart failure | 1462 (1.8) | 373 (6.7) | 1089 (1.4) | <.001 |
| Cerebrovascular disease | 3248 (3.9) | 491 (8.8) | 2757 (3.6) | <.001 |
| Ischemic heart disease | 8218 (10.0) | 3360 (60.0) | 4858 (6.3) | <.001 |
| High-risk surgery | 41396 (50.2) | 2775 (49.5) | 38621 (50.3) | .29 |
| Type of surgery |  |  |  | <.001 |
| General | 33652 (40.8) | 2380 (42.5) | 31272 (40.7) |  |
| Thoracic | 5959 (7.2) | 387 (6.9) | 5572 (7.3) |  |
| Transplant | 5566 (6.8) | 207 (3.7) | 5359 (7.0) |  |
| Vascular | 2345 (2.8) | 451 (8.0) | 1894 (2.5) |  |
| Urologic | 7779 (9.4) | 758 (13.5) | 7021 (9.1) |  |
| Breast and endocrine | 947 (1.1) | 67 (1.2) | 880 (1.1) |  |
| Neurosurgery | 11106 (13.5) | 427 (7.6) | 10679 (13.9) |  |
| Obstetrics and gynecology | 2361 (2.9) | 57 (1.0) | 2304 (3.0) |  |
| Orthopedic | 10849 (13.2) | 674 (12.0) | 10175 (13.2) |  |
| Otolaryngology | 1492 (1.8) | 136 (2.4) | 1356 (1.8) |  |
| Plastic | 385 (0.5) | 59 (1.1) | 326 (0.4) |  |
| Left ventricular ejection fraction |  |  |  |  |
| ≤ 40% | 712 (0.9) | 349 (6.2) | 363 (0.5) | <.001 |
| Not available | 7342 (8.9) | 218 (3.9) | 7124 (9.3) |  |
| Revised cardiac risk index |  |  |  | <.001 |
| 0 | 32498 (39.4) | 785 (14.0) | 31713 (41.3) |  |
| 1 | 39837 (48.3) | 2453 (43.8) | 37384 (48.7) |  |
| 2 | 8445 (10.2) | 1752 (31.3) | 6693 (8.7) |  |
| ≥3 | 1661 (2.0) | 613 (10.9) | 1048 (1.4) |  |
| Medication history* |  |  |  |  |
| Beta blocker | 16591 (20.1) | 2862 (51.1) | 13729 (17.9) | <.001 |
| Calcium channel blocker | 38279 (46.4) | 3693 (65.9) | 34586 (45.0) | <.001 |
| ACEi or ARB | 23904 (29.0) | 2608 (46.5) | 21296 (27.7) | <.001 |
| Statin | 19981 (24.2) | 3208 (57.3) | 16773 (21.8) | <.001 |
| Aspirin | 9594 (11.6) | 2684 (47.9) | 6910 (9.0) | <.001 |
| Clopidogrel | 4896 (5.9) | 1662 (29.7) | 3234 (4.2) | <.001 |

Data are presented as no. (%) of individuals unless otherwise indicated.
\*Medications at the time of admission for surgery.
Abbreviations: ACEi, angiotensin-converting enzyme inhibitor; ARB, angiotensin II receptor blocker; SD, standard deviation.

### Primary and Secondary Outcomes

At 30 days, 82388 (99.9%) of patients were completed clinical follow-up. The primary outcomes (the composite of cardiac death or myocardial infarction) occurred in 184 patients (97 cardiac deaths and 100 myocardial infarctions) within 30 days of elective noncardiac surgery. The causes of death are summarized in eTable 1. Figure 3 demonstrates that the cumulative incidences of the primary and secondary outcomes were all significantly higher among patients with abnormal MPI results than among those with normal results. As summarized in Table 2, the risk of the primary outcome was significantly higher in patients with abnormal MPI than in those with normal results (crude incidence, 1.2% vs 0.1%; adjusted OR, 4.64; 95% CI, 3.29 to 6.50; *P*<.001). Similarly, cardiac death (0.5% vs 0.1%; adjusted OR, 3.11; 95% CI, 1.86 to 5.07; *P*<.001), death from any cause (1.0% vs 0.5%; adjusted OR, 1.41; 95% CI, 1.03 to 1.89; *P*=.026), and myocardial infarction (0.9% vs 0.1%; adjusted OR, 8.19; 95% CI, 5.21 to 12.87; *P*<.001) were more frequent in patients with abnormal MPI. Among 23934 patients who underwent routine troponin testing after noncardiac surgery, the risk of MINS was also significantly higher in patients with abnormal MPI (16.5% vs 13.2%; adjusted OR, 1.37; 95% CI, 1.23 to 1.52; *P*<.001), as indicated in Table 2 and eFigure 1.

**Figure 3.**
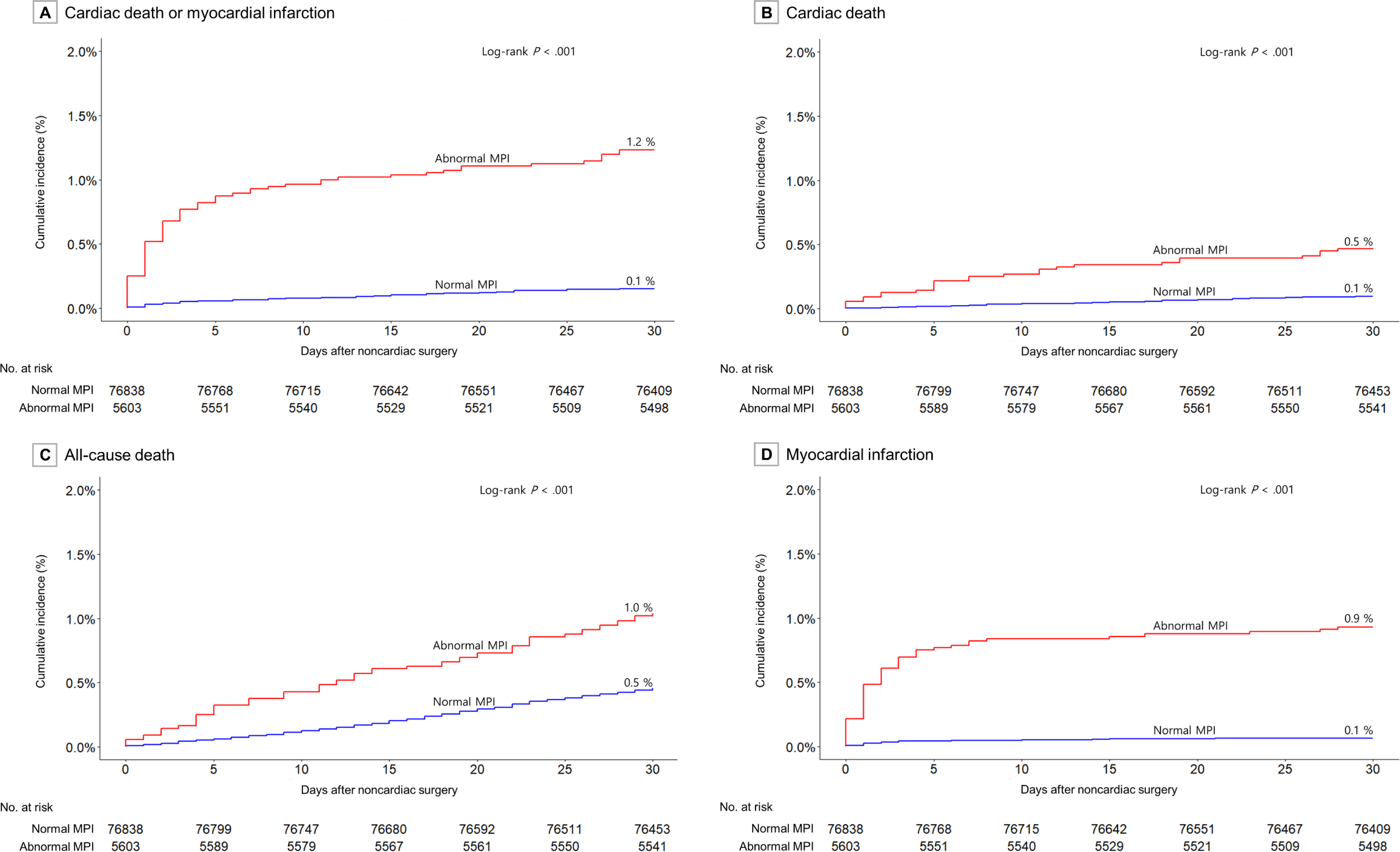
Primary and Secondary Outcomes. The cumulative incidences of the primary outcome of the composite of cardiac death and myocardial infarction (A) and the secondary outcomes of cardiac death (B), all-cause death (C), and myocardial infarction (D) within 30 days of noncardiac surgery. The *P* value was calculated by means of the log-rank test.

**Table 2.** Thirty-day Outcomes, stratified by MPI Results.

|  | Myocardial perfusion imaging<sup>a</sup> |  | Unadjusted |  | Adjusted<sup>b</sup> |  |
| --- | --- | --- | --- | --- | --- | --- |
|  | Abnormal | Normal | OR [95% CI] | *P* value | OR [95% CI] | *P* value |
|  | n=5598 | n=76790 |  |  |  |  |
| Primary outcome |  |  |  |  |  |  |
| Cardiac death or MI | 69 (1.2)<sup>a</sup> | 115 (0.1) | 8.32 [6.14-11.19] | <.001 | 4.64 [3.29-6.50] | <.001 |
| Secondary outcomes |  |  |  |  |  |  |
| Cardiac death | 26 (0.5) | 71 (0.1) | 5.04 [3.16-7.80] | <.001 | 3.11 [1.86-5.07] | <.001 |
| All-cause death | 58 (1.0) | 352 (0.5) | 2.27 [1.70-2.98] | <.001 | 1.41 [1.03-1.89] | .026 |
| MI | 52 (0.9) | 48 (0.1) | 15.0 [10.11-22.26] | <.001 | 8.19 [5.21-12.87] | <.001 |
| Patients undergoing troponin test<sup>c</sup> | n=3281 | n=20638 |  |  |  |  |
| Cardiac death and MINS | 548 (16.7) | 2741 (13.3) | 1.31 [1.18-1.45] | <.001 | 1.38 [1.24-1.53] | <.001 |
| MINS | 542 (16.5) | 2729 (13.2) | 1.30 [1.17-1.44] | <.001 | 1.37 [1.23-1.52] | <.001 |

Abbreviations: CI, confidence interval; OR, odds ratio; MI, myocardial infarction; MINS, myocardial injury after noncardiac surgery.
<sup>a</sup>Crude incidence within 30 days, no. of events (%).
<sup>b</sup>Adjusted variables were age, sex, revised cardiac risk index, and myocardial perfusion imaging result.
<sup>c</sup>Cardiac troponin test was conducted within 30 days after noncardiac surgery in 23934 patients, and 15 patients of them had missing data.

**Table 3.** Predictive Performance of Myocardial Perfusion Imaging before Noncardiac Surgery.

|  | AUC<sup>a</sup> | *P* value | Net reclassification improvement |  |  |  |
| --- | --- | --- | --- | --- | --- | --- |
|  |  |  | Events | Non-events | Overall [95% CI] | *P* value |
| Cardiac death and MI |  |  |  |  |  |  |
| Baseline model<sup>b</sup> | 0.73 |  |  |  |  |  |
| Plus MPI result | 0.77 | <.001 | -0.12 | 0.38 | 0.26 [0.11-0.40] | <.001 |
| Cardiac death |  |  |  |  |  |  |
| Baseline model<sup>b</sup> | 0.70 |  |  |  |  |  |
| Plus MPI result | 0.73 | .048 | -0.26 | 0.35 | 0.09 [-0.10-0.28] | .360 |
| All cause death |  |  |  |  |  |  |
| Baseline model<sup>b</sup> | 0.65 |  |  |  |  |  |
| Plus MPI result | 0.65 | .110 | -0.42 | 0.30 | -0.12 [-0.21-0.03] | .007 |
| Myocardial infarction |  |  |  |  |  |  |
| Baseline model<sup>b</sup> | 0.76 |  |  |  |  |  |
| Plus MPI result | 0.83 | <.001 | 0.10 | 0.49 | 0.59 [0.39-0.79] | <.001 |

Abbreviations: AUC, area under the receiver-operating-characteristic curve; CI, confidence interval; MPI, myocardial perfusion imaging.
<sup>a</sup>AUC for the relevant logistic regression model.
<sup>b</sup>Covariates in the baseline model were age, sex, and the revised cardiac risk index score.

When abnormal MPI findings were classified as fixed only or reversible, the primary outcome more frequently occurred among patients with a fixed defect only (adjusted OR, 3.42; 95% CI, 1.94 to 5.69; *P*<.001) and those with a reversible perfusion defect (adjusted OR, 5.26; 95% CI, 3.62 to 7.57; *P*<.001) than it did among patients with normal MPI, as demonstrated in eFigure 2A and eTable 2. In addition, the risk of the primary outcome increased according to the extent of ischemia. Compared with <5% ischemic burden (reference category), the adjusted OR for 5-10% ischemic burden was 1.47 (95% CI, 0.94 to 2.23; *P*=0.080), while >10% ischemic burden was associated with an adjusted OR of 3.52 (95% CI, 2.07 to 5.70; *P*<.001) (eFigure 2B and eTable 2).

### Subgroup Analysis

Figure 4 demonstrates the incidence of the primary and secondary outcomes according to the RCRI score. The risk of cardiac death or MI increased with increasing RCRI score. The prognostic impact of an abnormal MPI on the risk of cardiac death or MI was more prominent in patients with low RCRI risk category (*P* for interaction <.001). Nevertheless, even among the highest risk group (patients with RCRI ≥2 and abnormal MPI), the absolute risk of cardiac death or MI was only 1.5% at 30 days. Additional subgroup analyses according to the clinical subgroup and types of surgery, which consistently showed the higher risk of primary outcome in patients with abnormal MPI, were summarized in eFigure 3 and eFigure 4.

**Figure 4.**
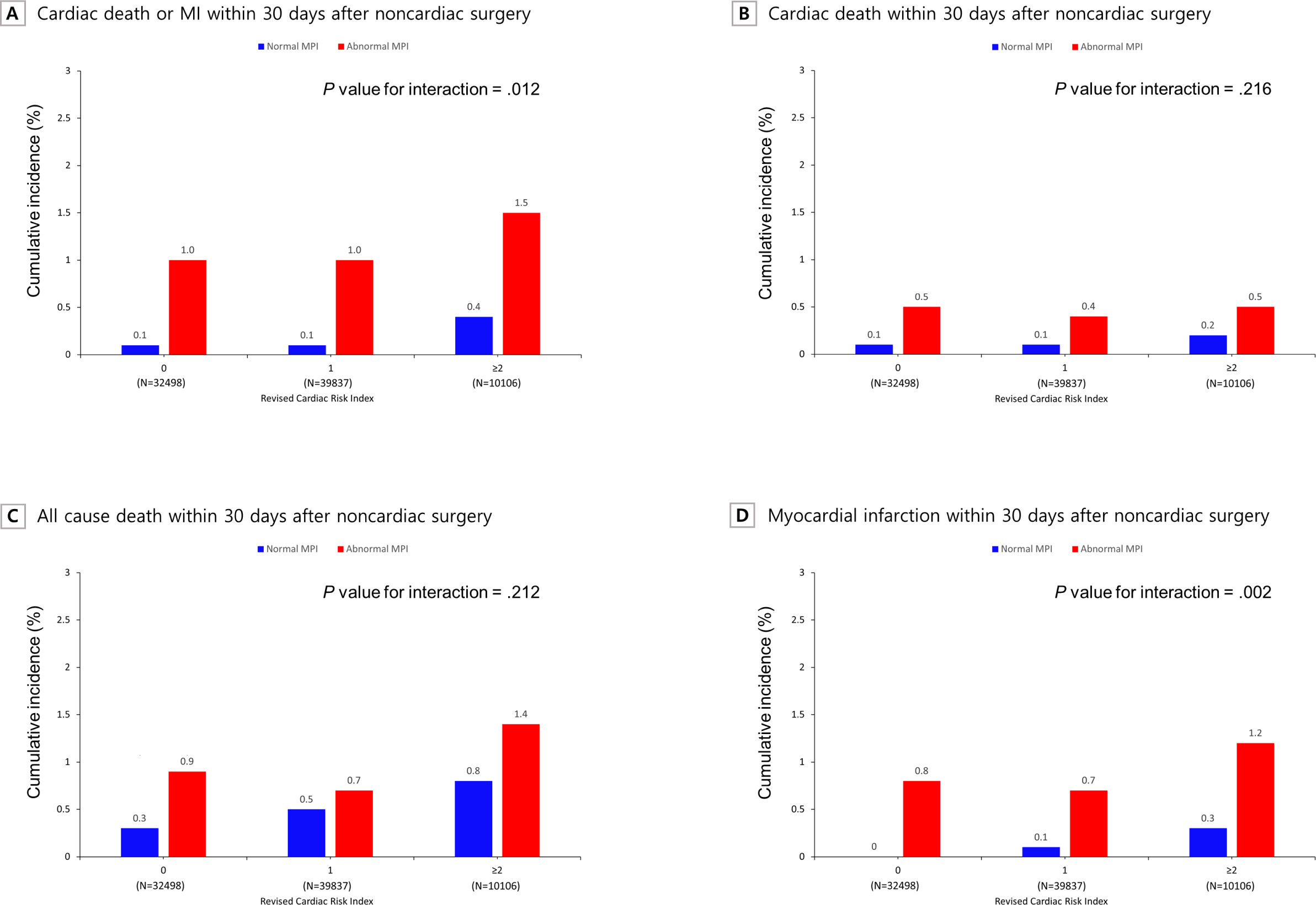
Primary and Secondary Outcomes According to Revised Cardiac Risk Index Score. Subgroup analyses by revised cardiac risk index score for primary and secondary outcomes, classified into three groups (score of 0, 1, and ≥2). The crude incidence of the outcomes is marked on the bars. P value for interaction is also shown.

### Prognostic Performance of MPI before Noncardiac Surgery

The presence of an abnormal MPI improved discrimination for the primary outcome (AUC with MPI vs without MPI [0.77 vs 0.73; *P*<0.001]) and significantly increased NRI (0.26, 95% CI, 0.11-0.40; *P*<0.001). These significant improvements were driven mainly by improved discrimination for myocardial infarction. The model including adjustment for abnormal MPI results yielded good discrimination performance for myocardial infarction (AUC=0.83).

### Coronary Angiography and Revascularization before Noncardiac Surgery

Among patients with abnormal MPI results (n=5603), 1743 underwent coronary angiography, and subsequently, 378 underwent coronary revascularization (260 percutaneous coronary interventions and 118 coronary artery bypass graft surgeries) before elective noncardiac surgery. Among patients with abnormal MPI, patient who underwent coronary angiography or revascularization were not significantly associated with the lower risk of the primary outcome within 30 days of noncardiac surgery (Figure 5).

**Figure 5.**
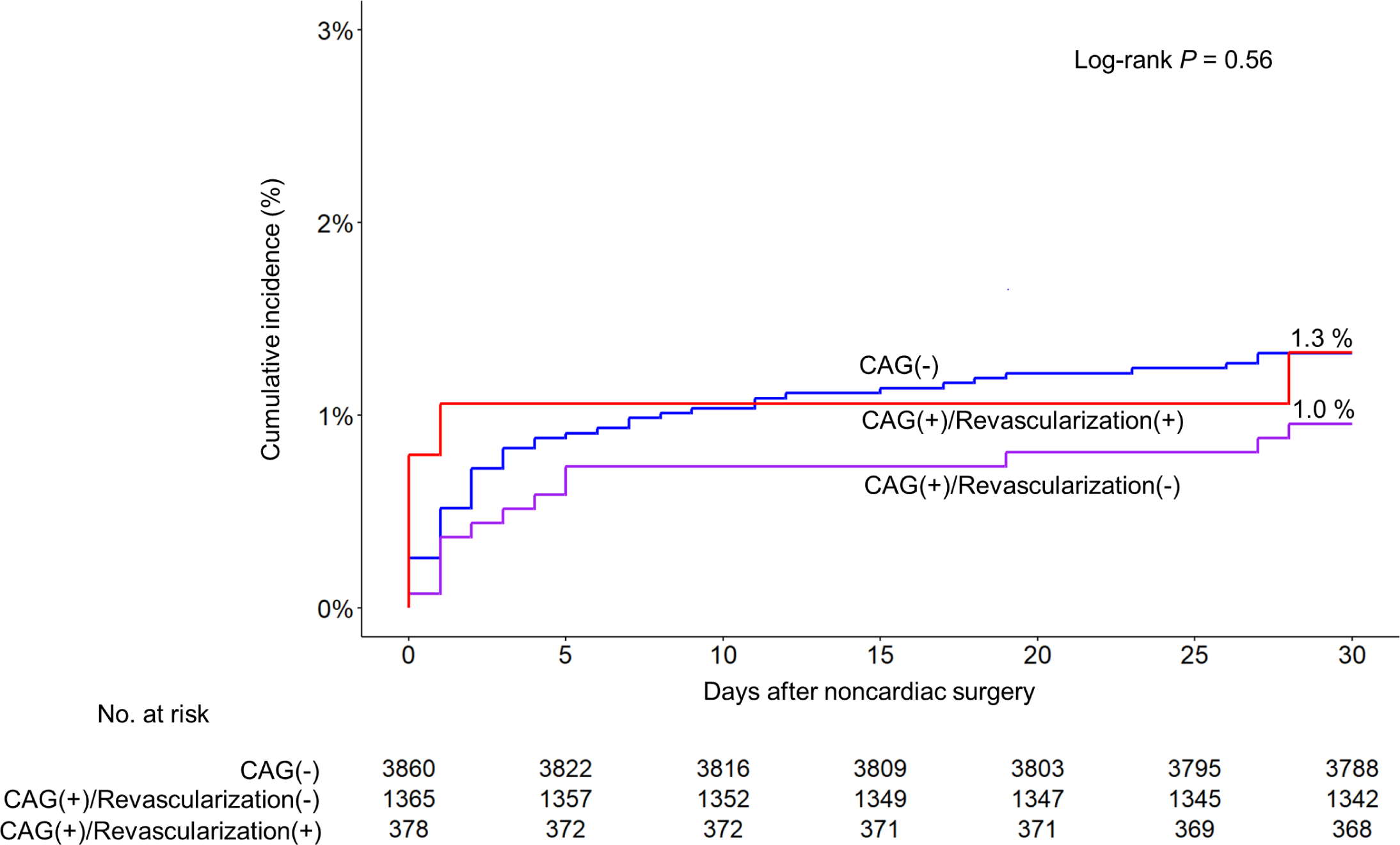
Primary Outcome According to Preoperative Coronary Angiography and Revascularization among Patients with Abnormal MPI Results. Cumulative incidences of the primary outcome of the composite of cardiac death and myocardial infarction within 30 days after noncardiac surgery. CAG, coronary angiography; MPI, myocardial perfusion imaging.

## Discussion

This large, observational study identified a significant association between an abnormal preoperative MPI and the composite of cardiac death or myocardial infarction within 30 days of noncardiac surgery—an association that increased progressively according to the extent of myocardial ischemia. When compared with standard clinical risk factors, the use of preoperative MPI testing led to a significant improvement in discrimination as well as substantial reclassification of risk as assessed by the net reclassification index. Nonetheless, given the low overall incidence of post-operative cardiac events in the study population, the positive predictive value of an abnormal MPI study to predict postoperative cardiac event was low (1.2%), leading to potentially unnecessary coronary angiography and revascularization procedures with unproven prognostic value.

Previous studies have examined a role for MPI in stratification of perioperative cardiac risk, but the results have been mixed.^5, 23–25^ A meta-analysis of nine studies including 1,179 patients undergoing noncardiac vascular surgery revealed that reversible defects in ≥20% of myocardial segments were significantly associated with perioperative complications.^5^ Another study suggested that incorporation of MPI may improve perioperative risk assessment of patients with obstructive disease upon coronary computed tomography angiography.^23^ However, other studies have shown that routine use of MPI before abdominal aortic surgery did not predict the risk of cardiac complications^26, 27^ and did not improve patient risk classification beyond essential assessment using age, RCRI, and surgical priority.^28^ However, These previous studies were limited by relatively small sample sizes and low event rates, inclusion of mainly patients undergoing relatively high-risk vascular surgery, and the use of outdated perfusion imaging techniques and perioperative management.^15, 29^ In contrast, our study included the largest population to date and included a broad spectrum of patients and surgical procedures across the full risk spectrum, thus demonstrating the consistent prognostic utility of MPI for predicting perioperative cardiac risk in contemporary practice.

Current clinical guidelines do not support the routine use of MPI in preoperative risk assessment and rather recommend a subjective assessment of functional capacity as an initial step for preoperative cardiac risk assessment.^9, 30^ Typically, preoperative MPI is recommended only for patients with both elevated risk of major adverse cardiac events and poor exercise capacity. However, a recent prospective cohort study has revealed that subjective assessments of functional capacity are neither an accurate predictors of exercise capacity based on formal cardiopulmonary testing nor associated with the risk of post-operative cardiac events.^22^ Moreover, recent studies suggested that the prevalence of poor functional capacity is relatively low in clinical practice, and the most patients who experience postoperative cardiac complications had satisfactory preoperative functional capacities.^31–33^ Therefore, our study suggested that more liberal use of MPI, ungated by functional capacity, is seemingly warranted, particularly in patients with a considerable surgical risk.

Prior to our study, there was little information on the value of preoperative non-invasive stress testing in low-risk patients.^24, 29^ Our study, which included 32498 patients with RCRI 0, demonstrated that even among low-risk patients, an abnormal MPI study was significantly associated with an increased risk of post-operative cardiac events. In addition, the prognostic impact of abnormal MPI was more prominent in patients with low cardiac risk. Nonetheless, given that the incidence of an abnormal MPI in low-risk patients (RCRI 0) was only 2.4% and the incidence of cardiac death or MI among these individuals was only 1.0%, the value of routine preoperative MPI testing in low-risk population should be interpreted in the context of the appropriate use of medical resources and cost-effectiveness in real practice.

The original justification for preoperative MPI was to identify patients with significant myocardial ischemia who would potentially benefit from coronary revascularization prior to noncardiac surgery. However, this hypothesis was refuted by the randomized trials which demonstrated no clinical benefit of coronary revascularization before noncardiac surgery^34–37^ – results that are reinforced by our observational data. Therefore, our study suggested that preoperative MPI should not be the sole indication for preoperative invasive coronary angiography and subsequent coronary revascularization in stable elective surgical candidates, which is supported by recent randomized trial in patients with stable ischemic heart disease.^38^ Nonetheless, previous observational study has suggested an association between performing preoperative MPI and reduced rate of perioperative mortality after noncardiac surgery.^29^ These findings suggest that clinical benefits associated with preoperative MPI may be explained by mechanisms other than coronary revascularization such as careful anesthesiologic care, meticulous perioperative medical surveillance and management, changes in surgical technique, mostly towards a less invasive approach, or even deferring surgery in some very high risk patients. Further prospective study is necessary to evaluate whether performing preoperative MPI and subsequent changes in surgical and medical management of patient would improve postoperative cardiac outcomes.

Our study has several limitations. First, it was a retrospective observational study. Therefore, the possibility of residual confounding in the associations cannot be eliminated despite the statistical adjustments we made for several key clinical characteristics. Second, our data sources did not capture information on exercise tolerance. Third, biomarkers for perioperative myocardial necrosis were not obtained for all patients. Fourth, the attending physicians and surgeons were not blinded to the MPI results, which might have affected the clinical outcomes. Fifth, this study was performed in a tertiary hospital with high surgical volumes, thus limiting its generalizability to lower-volume surgical centers.

In conclusion, among patients undergoing elective noncardiac surgery and referred for preoperative MPI testing, an abnormal MPI study was associated with an increased risk of 30-day cardiac death or MI—a result that was independent of age, sex, and clinical risk factors. However, the value of routine preoperative MPI testing appears to be limited given its low positive predictive value and the fact that coronary angiography or revascularization triggered by an abnormal MPI result was not associated with improved outcomes.

